# Liquid biomarkers of macrophage dysregulation and circulating spike protein illustrate the biological heterogeneity in patients with post-acute sequelae of COVID-19

**DOI:** 10.1101/2022.09.18.22280022

**Authors:** Christoph Schultheiß, Edith Willscher, Lisa Paschold, Cornelia Gottschick, Bianca Klee, Lidia Bosurgi, Jochen Dutzmann, Daniel Sedding, Thomas Frese, Matthias Girndt, Jessica I. Höll, Michael Gekle, Rafael Mikolajczyk, Mascha Binder

## Abstract

Post-acute sequelae of COVID-19 (PASC) are long-term consequences of SARS-CoV-2 infection that can substantially impair quality of life. Underlying mechanisms ranging from persistent virus to innate and adaptive immune dysregulation have been discussed. Here, we profiled plasma of 181 individuals from the cohort study for digital health research in Germany (DigiHero) including individuals after mild to moderate COVID-19 with or without PASC and uninfected controls. We focused on soluble factors related to monocyte/macrophage biology and on circulating SARS-CoV-2 spike (S1) protein as potential biomarker for persistent viral reservoirs. At a median time of eight months after infection, we found pronounced dysregulation in almost all tested soluble factors including both pro-inflammatory and pro-fibrotic cytokines. These perturbations were remarkably independent of ongoing symptoms, but further correlation and regression analyses suggested PASC specific patterns involving CCL2/MCP-1 and IL-8 as well as long-term persistence of high IL-5 and IL-17F levels. None of the analyzed factors correlated with the detectability or levels of circulating S1 indicating that this represents an independent subset of patients with PASC. This data confirms prior evidence of immune dysregulation and persistence of viral protein in PASC and illustrates its biological heterogeneity that still awaits correlation with clinically defined PASC subtypes.

## Introduction

The coronavirus disease 2019 (COVID-19) caused by the zoonotic severe acute respiratory syndrome coronavirus 2 (SARS-CoV-2) is a systemic multi-organ disease with a broad severity spectrum ranging from asymptomatic to fatal outcomes, especially in risk groups [1, 2]. While most individuals mount lasting SARS-CoV-2-directed immune responses [3] and the rapid development of effective and safe vaccines helped to prevent severe disease courses and mitigate the pandemic progression, it is now clear that a substantial proportion of SARS-CoV-2 infected individuals does not fully recover but has persisting health impairments beyond four weeks of symptom onset that can last for months and significantly impact the quality of life [4, 5]. These post-acute sequelae of COVID-19 (PASC), or post-COVID-19 condition as suggested by the WHO, are reported in 12.7-87% of patients and encompass a wide range of systemic, respiratory, neuropsychiatric and cardiac manifestations including fatigue, head and body aches, memory defects, dyspnea, palpitations as well as sleep and anxiety disorders [4-9]. Preexisting comorbidities like obesity and diabetes as well as age and severity of acute disease might represent risk factors, but lasting symptoms are also common among young individuals with mild disease courses and after vaccination [10, 11].

While the epidemiological and clinical characterization of PASC is relatively advanced, mechanistic insights in the pathophysiological underpinnings of this condition are still limited. A potential trigger of ongoing sequelae are persistent immunogenic viral reservoirs. SARS-CoV-2 RNA and spike or other proteins that might fuel ongoing and generate de-novo SARS-CoV-2-specific or superantigenic T cell responses [10, 12-14] have been detected in the respiratory tract, the gut, the brain, kidney and circulating in the blood months after acute disease [1, 15-17]. These findings might also mirror unrepaired virus-induced tissue damage that could account for some of the organ-specific symptoms in PASC [5, 18-20]. Autoimmunity represents another potential driver of PASC. Adaptive immune responses during acute COVID-19 show imprints of autoreactivity and are characterized by the production of a variety of different autoantibodies that are also found in post-acute phases and in the SARS-CoV-2-induced post-infection multisystem inflammatory syndrome in children (MIS-C) [6, 21-24]. In addition, dysbiosis of the microbiome that either results in the persisting production of inflammatory mediators like LPS that might promote inflammation or the long-term depletion of anti-inflammatory modulators is discussed [25, 26].

We recently reported persisting elevation of a triad of monocyte/macrophage-related cytokines - IL-1β, IL-6 and TNF 8-10 months after SARS-CoV-2 infection in patients with PASC [6]. We hypothesized that these cytokines are secreted by tissue-resident macrophages that engage into a self-sustaining proinflammatory loop that may fuel PASC. Such macrophage imprinting has been previously reported to be potentially induced through engulfment of spike protein by tissue-resident macrophage in early disease phases [27, 28].

In order to obtain a broader picture of the profiles of immune dysregulation, their variability across patients and their relation to persisting virus or viral antigen, we designed a refined liquid biomarker panel to be run on biosamples from the DigiHero study cohort. Our data illustrate the pronounced dysregulation of monocyte/macrophage-related soluble factors in some individuals with PASC and the long-term circulation of spike protein in others. Together, this data further refines the molecular underpinnings of PASC and suggests the existence of different PASC subtypes.

## Materials and methods

### The population-based cohort study for digital health research in Germany (DigiHero)

The here analyzed individuals essentially reflect the discovery cohort of the COVID-19 module of the DigiHero study [6]. This subcohort encompasses 181 participants from the DigiHero discovery cohort who were interviewed with an online questionnaire on the clinical course of their SARS-CoV-2 infection, its post-infection sequelae and SARS-CoV-2 vaccination status. Interviews and blood sampling was performed until 9^th^ of October 2021. The study was approved by the institutional review board (approval numbers 2020-076) and conducted in accordance with the ethical principles stated by the Declaration of Helsinki. Informed written consent was obtained from all participants or legal representatives. Plasma samples were isolated by centrifugation of whole blood for 15 minutes at 2000 x g, followed by centrifugation at 12000 x g for 10 minutes. Samples were stored at - 80°C until further use.

### Biological samples and data from the biobank of the Halle COVID cohort (HACO)

Plasma samples from acute COVID-19 (n=15 mild to moderate severity) were used as control group. Samples were collected between April and December 2020. Informed written consent was obtained and the study was approved by the institutional review board (approval number 2020-039) and conducted in accordance with the ethical principles stated by the Declaration of Helsinki. The collected plasma samples were isolated as described above.

### Profiling of human plasma for monocyte/macrophage-related soluble factors, anti-SARS-CoV-2 antibodies and circulating SARS-CoV-2 spike protein

Plasma levels of IL-5, IL-9, IL-17F, IL-18, IL-22, IL-23, IL-33 and CCL2/MCP-1 were measured using the respective LEGENDplex capture beads and corresponding detection antibodies from the LEGENDplex Human Inflammation Panel and Human Th Panel (BioLegend). For quantification of soluble CD206 (MMR), the Human MMR ELISA (RayBiotech) was used, for quantification of soluble CD163 the Human CD163 Quantikine ELISA Kit (R&D Systems). Profiling of antibodies directed against the spike (S1) protein and the nucleocapsid protein (NCP) of SARS-CoV-2 was performed using the anti-SARS-CoV-2-ELISA IgG and anti-SARS-CoV-2-NCP-ELISA kits from Euroimmun (Lübeck, Germany). Circulating S1 protein was measured using the RayBio COVID-19 S-Protein (S1RBD) ELISA kit (RayBiotech). All kits were used according to the manufacturer’s instructions. Read out of the LEGENDplex system was performed on a BD FACSCelesta, ELISAs were read on a Tecan Spark Microplate reader.

### Statistical analysis

All bar/dot plots as well as logistic regression and Spearman rank-order correlation analysis for plasma levels over time were generated using GraphPad PRISM 8.3.1 (GraphPad Software, La Jolla, CA, USA). Differences in plasma cytokine levels were studied by unpaired t-test with Welch’s correction and Welch’s ANOVA. Correlations were calculated using the R package corrplot. Ranges of p values are indicated with asterisks: *p<0.05; ***p<0.01; are indicated with asterisks ****p<0.001; are indicated with asterisks p<0.0001.

## Results

### Characteristics of post-acute COVID-19 cohort

To study post-COVID-19 perturbations in monocyte/macrophage-related soluble factors, we randomly selected 181 individuals from the discovery cohort of DigiHero [6] for profiling. The subcohort consisted of 91 individuals with ongoing PASC at the time of blood sampling (65 females, 26 males), 62 individuals who never reported PASC (26 females, 36 males) and 28 individuals without prior COVID-19 (17 females, 11 males) (Figure 1A). All participants were recruited until October 2021. Median time from infection to sampling was 8 months for individuals with ongoing PASC (range 1-17 months) and 7.5 months for individuals without PASC (range 4-17 months) (Figure 1B), median age was 51 for both post-COVID-19 groups and 50 for the never COVID-19 group (Figure 1C). Most individuals of the analyzed groups had received at least one vaccination (73% of individuals without PASC, 77% of individuals with ongoing PASC, 83% of individuals without prior COVID-19).

**Figure 1.**
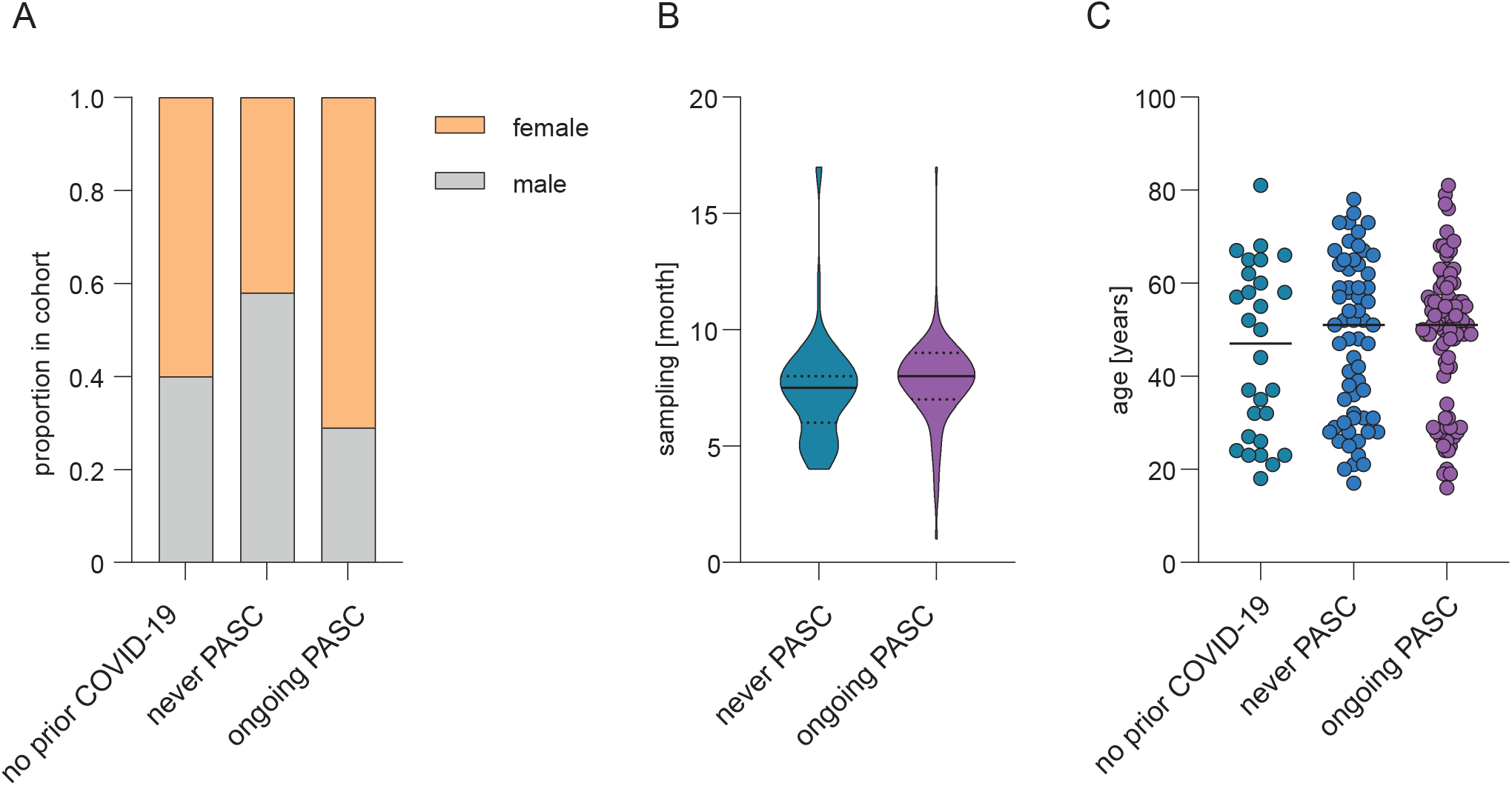
Clinical and epidemiological characteristics 384 of the post-COVID-19 cohort. **(A)** Sex distribution in the analyzed cohort comprising individuals with ongoing PASC (n=91), individuals with prior COVID-19 who never reported PASC (n=62) and individuals without prior COVID-19 (n=28). **(B)** Violin plot of median blood sampling time point (continuous line) relative to positive PCR or antigen test for the post-COVID-19 groups. Dotted lines separate quartiles. **(C)** Median age of indicated groups.

### Plasma soluble factors associated with pro-inflammatory and pro-fibrotic macrophages are increased in post-acute COVID-19

We selected soluble plasma factors for profiling that are associated with distinct activation states or phenotypes of monocytes/macrophages in COVID-19. Our panel included both pro-inflammatory cytokines (e.g., IL-17, IL-18, IL-23) and more pro-fibrotic cytokines (e.g., IL-5, IL-9) [29-31]. We also included the shedded forms of two characteristic monocyte/macrophage surface molecules, namely the soluble mannose receptor (sMMR/sCD206/sMRC1) and the soluble haptoglobin-hemoglobin receptor (sCD163) [32].

We observed markedly increased plasma levels of IL-5, IL-9, IL-17F, IL-18, IL-22, IL-23, IL-33, CCL2/MCP-1 and sCD163 but only marginally increased levels for sCD206/sMMR in post-COVID-19 disease phases as compared to individuals who never had COVID-19 (Figure 2). The mean levels of IL-5, IL-9, IL-17F, IL-22, IL-23 and IL-33 tended towards higher values in individuals with ongoing PASC as compared to individuals who never reported PASC, while this trend was reversed for IL-18 and CCL2/MCP-1 (Figure 2).

**Figure 2.**
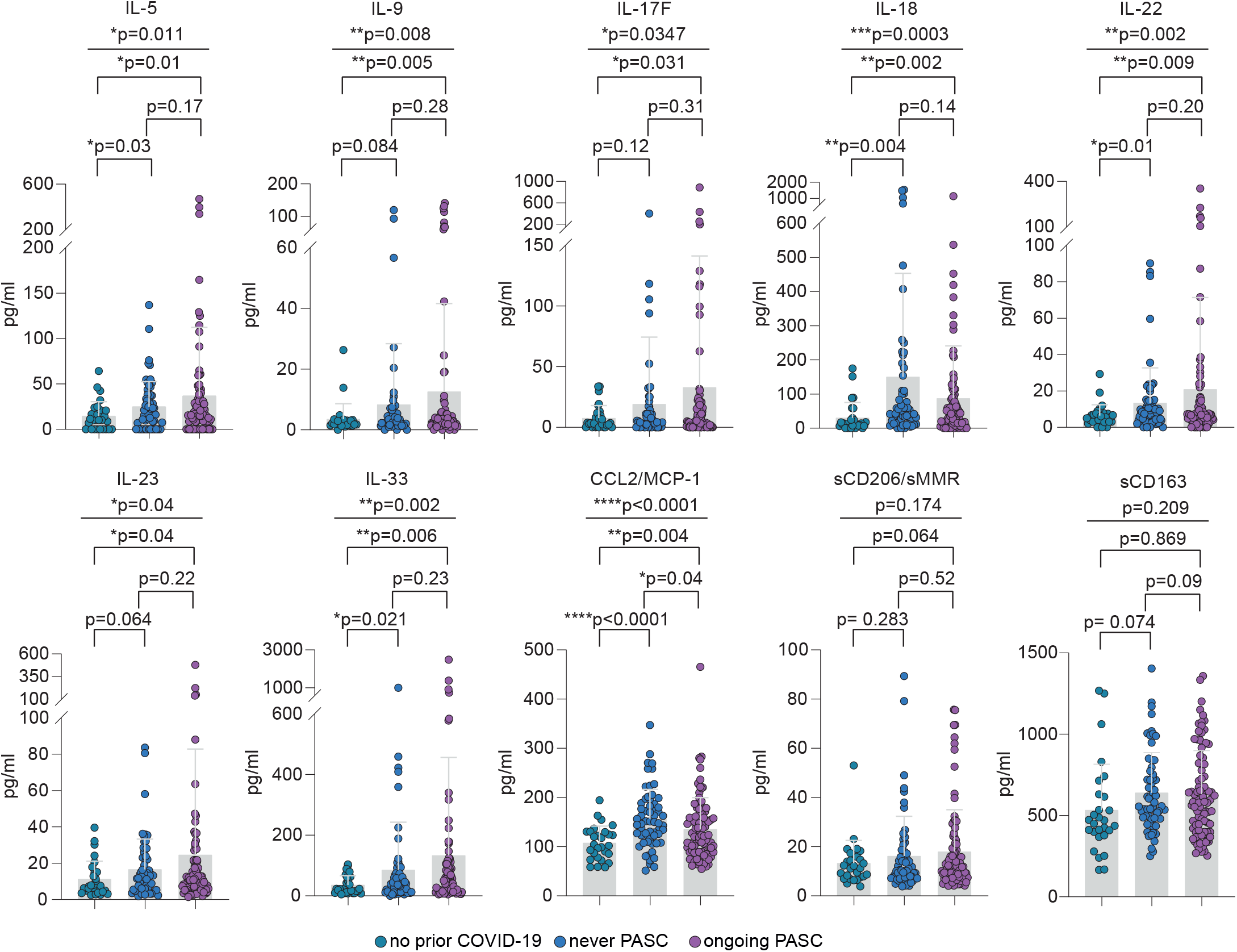
Profiling of plasma monocyte/macrophage-related soluble factors from individuals with ongoing PASC, without PASC and without SARS-COV-2 infection. Mean plasma cytokine/chemokine/soluble factor levels of individuals with no prior COVID-19 (n=28), individuals who never reported PASC post-infection (n = 62) or with ongoing PASC (n = 91). Error bars indicate ± SD. Statistical analysis: Welch’s ANOVA for comparison of all three groups and two-sided Welch corrected t-test for comparison of the no prior COVID-19 vs never PASC, no prior COVID-19 vs ongoing PASC and never PASC vs ongoing PASC groups.

To explore potential clusters of dysregulated soluble factors, we performed a large correlation analysis also including IL-1β, IL-4, IL-6, IL-8, IL-13, IL-17A, TNF, LTA (TNF-β) and IFN-α2 from our previously published report since these factors have been found increased in post-infection biosamples [6]. This analysis revealed a characteristic pattern of correlating cytokines in post-COVID-19 samples relatively independent of PASC (Figure 3A-B), but not in uninfected individuals (Figure 3C). There were only very few significant correlations that were evident in the PASC setting, but not in post-COVID-19 patients without PASC. Two factors that were remarkable in this respect were CCL2/MCP-1 and IL-8. Both showed no specific correlations in individuals without prior COVID-19 or without PASC, but a clear positive correlation with each other and additional factors in individuals with PASC (Figure 3A-C). CCL2/MCP-1 was strongly correlated with IL-18 and IL-23; IL-8 was correlated with the shedded macrophage molecules sCD206/MMR and sCD163 as well as IL-17A, IFN-α2 and IL-33. This pointed at potential PASC subgroups.

**Figure 3.**
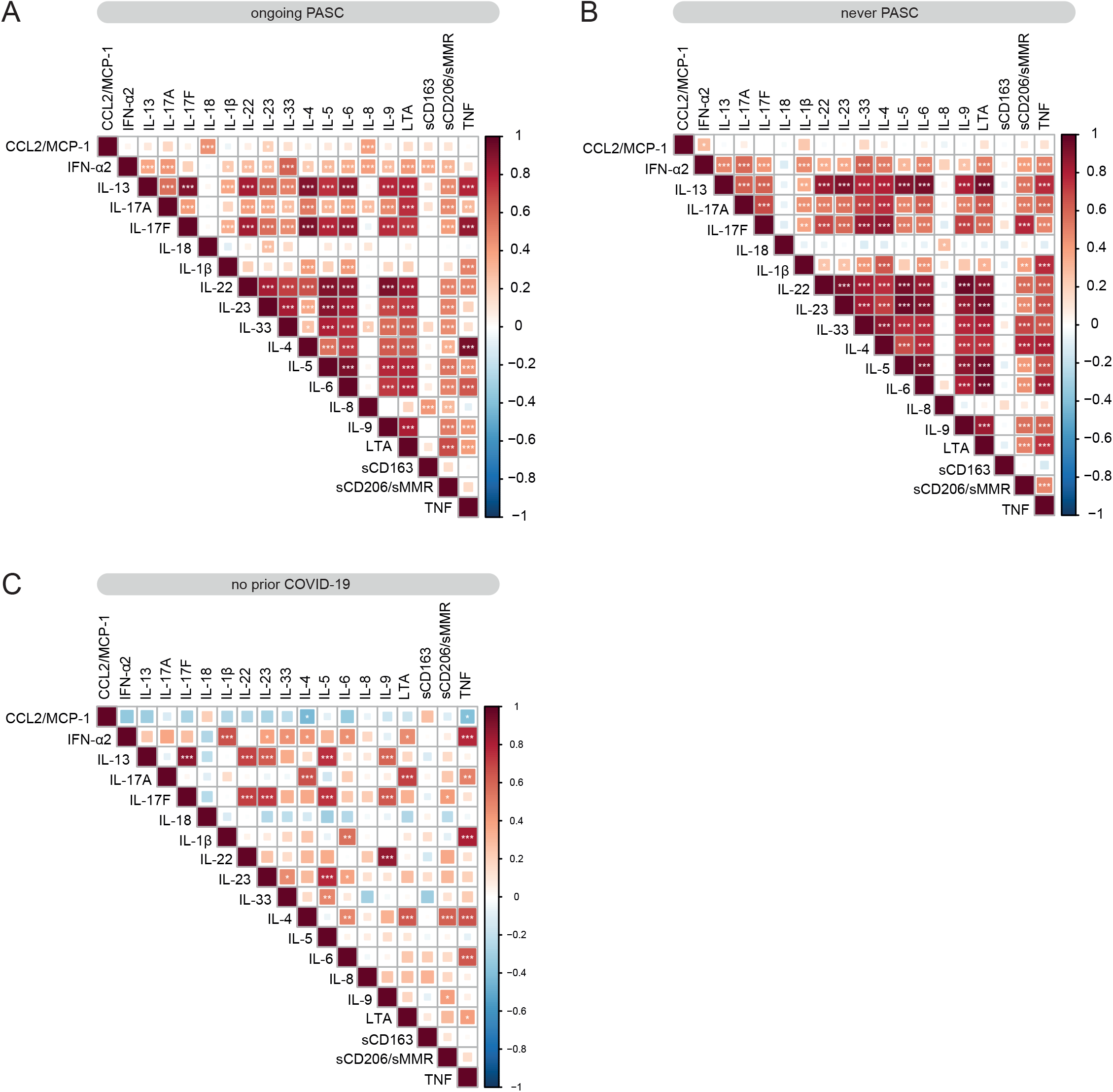
Correlation analysis of plasma soluble factors. **(A)-(C)** Correlation matrix of all analyzed soluble factors for individuals with ongoing PASC (n = 91) **(A)**, individuals who never reported PASC post-infection (n = 62) **(B)** or individuals with no prior COVID-19 (n=28) **(C)**.

### Some perturbations persist longer in patients with PASC compared to individuals without PASC

Next, we asked if individuals with PASC showed slower normalization of the strong perturbations in monocyte/macrophage-related factors than individuals without PASC. Since no repetitive sampling was performed in the DigiHero cohort, this analysis was restricted to interpatient comparisons (Figure 1B). In order to close the gap of early post-infection samples that were unavailable in the DigiHero cohort, we quantified the set of soluble factors in additional plasma samples from individuals with mild to moderate acute COVID-19 (nine females, six males; median age 68 [range 23-85]; median sampling on day 15 after symptom onset [range 1-23]) collected as part of the independent Halle COVID-19 (HACO) cohort [33]. Linear regression and Spearman rank-order correlation analysis revealed a relatively clear negative correlation between sampling time point and plasma levels for many of the dysregulated soluble factors in individuals without PASC (Figure 4A). In patients with PASC, such over-time normalization was less evident for some of the measured factors (Figure 4B). While Spearman correlation analysis showed a quite clear negative time correlation for IL-5 and IL-17F in individuals without PASC, this was not observed in individuals with PASC (Figure 4B). This suggested that individuals with PASC may show prolonged perturbations of the analyzed soluble factors.

**Figure 4.**
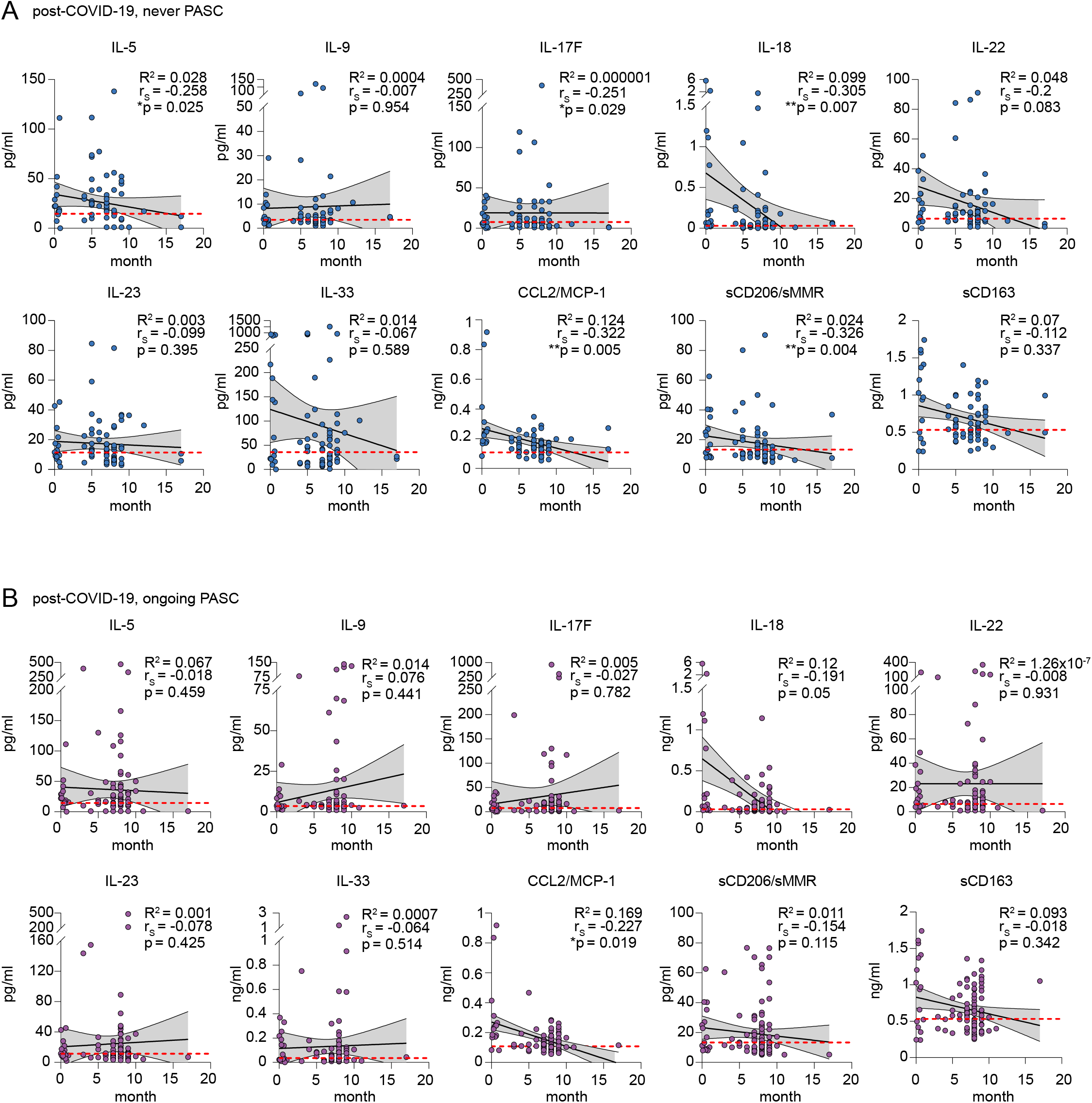
Association of plasma soluble factors with sampling time point post infection. **(A)-(B)** Linear regression of plasma cytokine levels and sampling time point post-infection in individuals without PASC (n =62) **(A)** and with ongoing PASC (n=91) **(B)**. Both cohorts also comprise cytokine data from individuals with mild/moderate acute COVID-19 (n=15). Dotted red lines indicate mean plasma level determined in individuals without prior COVID-19 (n=28). Correlation coefficient R_2_, Spearman correlation coefficients (r_S_) and p values are indicated.

### Plasma levels of circulating spike protein are detectable in a substantial proportion of patients after COVID-19 especially in those with PASC

Persistent immunogenic viral antigens like the SARS-CoV-2 S1 spike protein are potential drivers of PASC that might also fuel systemic cytokine and chemokine perturbations. To study this hypothesis, we profiled our cohort for levels of circulating S1. Since the S1 antigen has been detected in plasma after vaccination [34, 35], we restricted this analysis to individuals without prior vaccination. Around 35% of individuals with prior COVID-19 but no PASC showed measurable levels of circulating S1 protein (Figure 5A). In the ongoing PASC group, circulating S1 was detected in around 64% of individuals (Figure 5A). This group also showed numerically higher circulating S1 levels as compared to individuals without PASC (Figure 5B). However, the detectability or level of circulating S1 did not show a clear correlation with any of the soluble factors dysregulated in individuals with ongoing PASC (Figure 5C). Nevertheless, it should be noted that for three individuals with detectable plasma S1 the levels of TNF, IL-1β, IL-6 and/or IL-8 were in the upper range of values detected in the ongoing PASC group. Of note, levels of circulating S1 showed a trend towards positive correlation with S1 and NCP antibody levels suggesting that persistent viral proteins may sustain the immune response (Figure 5C).

**Figure 5.**
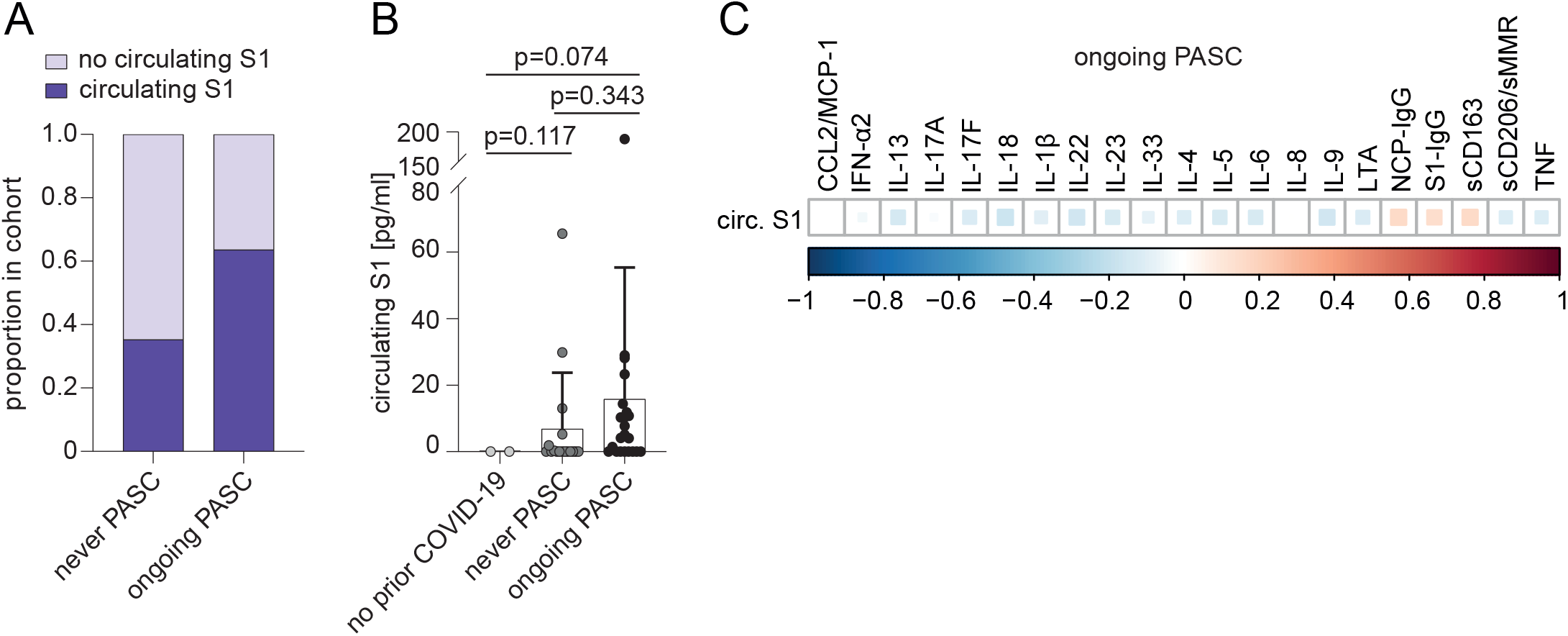
Persistence of circulating 418 S1 protein and correlation with monocyte/macrophage-related soluble factors and SARS-CoV-2-directed antibodies in unvaccinated individuals with ongoing PASC. **(A)** Proportion of individuals with detectable levels of circulating S1 (cS1) protein in unvaccinated individuals with prior COVID-19 who never experienced PASC (n=17) and with ongoing PASC (n=22). **(B)** Mean plasma levels of circulating SARS-CoV-2 spike (S1) protein in unvaccinated individuals with ongoing PASC (n=22), individuals with prior COVID-19 who never reported PASC (n=17) and individuals without prior COVID-19 (n=2). Error bars indicate ± SD. Statistical analysis: one-sided Welch corrected t-test. **(C)** Correlation matrix of indicated soluble factors with levels of circulating S1 and S1/NCP antibodies in unvaccinated individuals with ongoing PASC.

## Discussion

In this work, we provide evidence for the long-term and surprisingly strong dysregulation of monocyte/macrophage-related cytokines, chemokines and other soluble factors in individuals with a history of COVID-19. While individuals with PASC tended to show more dysregulation, the correlation patterns of these factors were remarkably independent of ongoing symptoms with a few exceptions. We also observed circulating SARS-CoV-2 S1 spike protein in a substantial proportion of individuals with a history of COVID-19 even many months after infection – especially in the subset of individuals with PASC. While the soluble “immune” factors showed strong correlations with each other, we did not find a strong correlation with the detectability or level of circulating S1. This was a relevant finding that we interpreted as indicative of distinct subgroups of PASC that may result from divergent underlying mechanisms. Unfortunately, the presumable PASC subsets suggested by our analyzes – individuals with predominant macrophage dysregulation versus individuals with persistent viral proteins or reservoirs – were rather small subgroups. Therefore, no reliable correlation analysis of these molecular patterns with the clinical characteristics registered in the context of the DigiHero trial could be performed.

Our screening effort in this well characterized cohort of patients focused strongly on the monocyte/macrophage compartment and its network of soluble factors. Monocytes and macrophage represent one of the most important cellular immune subsets that is associated with the heterogeneous courses and severity of acute COVID-19 [36, 37] and is also discussed as central for PASC [14, 27, 28, 38-41]. The here reported cytokine and chemokine data not only corroborates this hypothesis and the importance of pro-inflammatory and pro-fibrotic monocytes and macrophages [6], it also suggests a complex role of monocyte/macrophage-centered factors known to regulate the T_H_1/T_H_2 balance in PASC. This is in line with the reported differential activation of classical and non-classical monocytes in PASC [14, 42]. One of the most emblematic cytokines in this respect is IL-33, which has been originally described as a pro-inflammatory member of the IL-1 family but can also induce T_H_2 responses and act as damage-associated molecular pattern (DAMP). IL-33 was suggested to drive acute severity of COVID-19 in concert with GM-CSF, to mediate T_H_2 polarization and induce chronic pulmonary fibrosis. In addition, it may also mediate differentiation of monocytes to alternatively activated macrophage that may regenerate damaged bronchial epithelial tissue [43, 44].

An evolving body of evidence suggests that the wide spectrum of PASC symptoms mirrors the existence of different pathological subgroups [42, 45, 46]. In line with this notion, we identified two PASC-specific correlation patterns consisting of IL-8 and CCL2/MCP-1 with either sCD162, sCD206/MMR, IFN-α2, IL-17A and IL-33, or IL-18 and IL-23 which might hint towards distinct disease mechanisms. The correlation of IL-17A with IFN-α2 and IL-8 is notable given the importance of type I interferons for SARS-CoV-2 clearance and the pathogenic role of tissue-resident T_H_17 cells that interact with pro-inflammatory and pro-fibrotic macrophages in the lung of SARS-CoV-2-infected individuals leading to IL-8 secretion [47]. In contrast, the correlation of CCL2/MCP-1, IL-8, IL-18 and IL-23 in a subset of participants might be interpreted as a transition from pro-inflammatory T_H_1-like responses in the acute phase towards a more pronounced T_H_2 response in PASC that is associated with macrophage-dependent lung fibrosis potentially driven by an exacerbated reaction to type 2 cytokines. This is also in line with the lower levels of CCL2/MCP-1 and IL-8 in individuals with ongoing PASC as compared to individuals without PASC after SARS-CoV-2 infection that was also observed by others [40]. CCL2/MCP-1, IL-8, IL-18 and IL-23 have all been described as pro-fibrotic in lung, liver and/or heart [48-51] and might indicate ongoing tissue damage in PASC [52, 53]. Interestingly, IL-8 was found in all PASC-associated cytokine signatures underscoring its importance for long-lasting sequelae.

Persistent viral antigens, especially the S1 spike and NCP proteins, have been detected in multiple tissues post-infection and might provide a reservoir sustaining immune responses [1, 15, 16, 54]. We also observed persisting circulating S1 in post-infection samples with higher frequency of detection and increased levels in individuals with ongoing PASC. Notably, S1 levels did numerically correlate with SARS-CoV-2 antibody titers but not with any of the analyzed soluble immune factors. Nevertheless, a few individuals with circulating S1 had relatively high plasma levels of TNF, IL-1β, IL-6 and/or IL-8 supporting the superantigenic features of the spike protein [13]. In line with a recent publication analyzing 31 PASC patients [17], this data suggests that individuals with PASC that have circulating S1 represent a different disease subset independent of monocyte/macrophage reprogramming. In addition, there are individuals with circulating S1 post-infection who do not develop PASC. Given the small number of involved samples in both data sets, the pathological relevance of circulating S1 needs further validation in larger cohorts.

Overall, these data are indicative of a variety of molecular subtypes in PASC that need to be dissected in future studies with a clear theragnostic aim.

## Data Availability

All data produced in the present work are contained in the manuscript.

## Acknowledgement

The DigiHero study is conducted by a consortium of the Medical Faculty of the Martin-Luther-University Halle-Wittenberg including the following PIs: Mascha Binder, Thomas Frese, Michael Gekle, Matthias Girndt, Jessica I. Höll, Patrick Michl, Rafael Mikolajczyk, Matthias Richter and Daniel Sedding. We thank Aline Patzschke, Christoph Wosiek, Katrin Nerger and Bianca Gebhardt for excellent technical assistance. We sincerely thank healthy donors, patients and their household members for participating in this study. Flow cytometry was performed at the UKH FACS sorting core facility. This project was partially funded by the CRC 841 of the German Research Foundation (to MB) as well as by the Medical Faculty of the Martin-Luther-University Halle (Saale).

## Author contributions

M. Binder, R. Mikolajczyk, M. Gekle, C. Schultheiß, L. Paschold, C. Gottschick, B. Klee, M. Girndt, T. Frese, D. Sedding and J. I. Höll designed the COVID-19 module of the DigiHero cohort study. D. Sedding and J. Dutzmann provided the HACO patient cohort. C. Schultheiß and L. Paschold conducted experiments. M. Binder, C. Schultheiß, L. Paschold, E. Willscher and L. Bosurgi analyzed and interpreted the data. M. Binder, C. Schultheiß, E. Willscher and L. Paschold drafted the manuscript. All authors critically revised and approved the manuscript.

